# Cervical cancer screening uptake in Sub-Saharan Africa: a systematic review and meta-analysis

**DOI:** 10.1101/2020.12.26.20248864

**Authors:** Nigus Bililign Yimer, Mohammed Akibu Mohammed, Kalkidan Solomon, Mesfin Tadese, Stephanie Grutzmacher, Henok Kumsa Meikena, Birhan Alemnew, Nigussie Tadesse Sharew, Tesfa Dejenie Habtewold

## Abstract

**Background:** Cervical cancer screening and prevention programs have been given considerable attention in high-income countries, while only receiving minimal effort in many African countries. This meta-analytic review aimed to estimate the pooled uptake of cervical cancer screening uptake and identify its predictors in Sub-Saharan Africa.

**Methods:** PubMed, EMBASE, CINAHL, African Journals Online, Web of Science and SCOPUS electronic databases were searched. All observational studies conducted in Sub-Saharan Africa and published in English language from January 2000 to 2019 were included. The Newcastle-Ottawa Scale was applied to examine methodological quality of the studies. Inverse variance-weighted random-effects model meta-analysis was done to estimate the pooled uptake and odds ratio of predictors with 95% confidence interval. I^2^ test statistic was used to check between-study heterogeneity, and funnel plot and Egger’s regression statistical test were used to check publication bias. To examine the source of heterogeneity, subgroup analysis based on sample size, publication year and geographic distribution of the studies was carried out.

**Results:** Of 3,537 studies identified, 29 studies were included with 36,374 women. The uptake of cervical cancer screening in Sub-Saharan Africa was 12.87% (95% CI: 10.20, 15.54; I^2^= 98.5%). Meta-analysis of seven studies showed that knowledge about cervical cancer increased screening uptake by nearly 5-folds (OR: 4.81; 95% CI: 3.06, 7.54). Other predictors include educational status, age, HIV status, contraceptive use, perceived susceptibility, and awareness about screening locations.

**Conclusion:** Cervical screening uptake is low in Sub-Saharan Africa and influenced by several factors. Health outreach and promotion targeting identified predictors are needed to increase uptake of screening service in the region.s

**Protocol registration:** CRD42017079375

## Introduction

To date, cervical cancer is one of the global public health challenge (1). The primary cause of cervical pre-cancer and cancer is persistent infection with one or more of the high-risk oncogenic types of human papillomavirus (HPV) which interferes with the normal functioning of cells that results in distinct changes in the epithelial cells of transformation zone of the cervix (2). Cervical cancer is one of the very few type of cancers where a pre-cancer stage lasts many years before becoming invasive cancer that provide ample opportunity for detection and treatment (3). Cervical cancer is a malignancy for which screening is available. The screening seeks to identify pre-cancerous cellular changes on the cervix that may become cervical cancer if they are not appropriately treated (4).

Cervical cancer is the fourth most common cancer in women, with an estimated 530,000 new cases every year, representing 7.9% of all female cancers (5). In 2015, approximately 90% of the 270,000 deaths from cervical cancer occurred in low- and middle-income countries (5). Mortality rate remarkably varies among different regions of the world, with rates ranging from less than 2 per 100,000 in Western Europe and New Zealand to 27.6 per 100,000 in Sub-Saharan Africa (6).

Cervical cancer prevention and impact of screening program on cervical cancer related deaths has been given a considerable attention in developed countries with much minimal effort in most low and middle income nations (7). Cervical cancer screening coverage is very limited in low- and middle-income countries, as shown by a study which reported coverage of cervical cancer screening in developing countries on average to be 19% compared to 63% in developed countries (8). Data from the <<year>> World Health Survey indicated that the coverage of cervical cancer screening was 10% in Sub-Saharan Africa (9). Likewise, less than 1% of women in four West African countries had ever been screened for cervical cancer (10).

Even though cervical cancer screening is proven to reduce cervical cancer incidence, many factors influence screening uptake (11). Women’s rates of screening uptake have been shown to vary by knowledge about cervical cancer, and screening services, and other factors such as individual perception, beliefs, attitudes, and culture; and partner attitude (12). Several studies suggested that many women, particularly those with low levels of knowledge about cervical cancer and screening, may not recognize the benefit of screening over the possible consequences of forgoing screening (13-18).

Though it is very limited in scope, there are prevention, treatment, and rehabilitation strategies for cervical cancer such as risk assessment, screening, and clinical interventions in Sub-Saharan Africa. Nevertheless, they are not being fully utilized because of structural and behavioral barriers (19, 20). In order to enhance cervical cancer screening and treatment efforts, it is necessary to identify the factors affecting eligible women’s screening uptake and their prevalence. In this meta-analytic review, therefore, we aimed to estimate the pooled prevalence of cervical cancer screening uptake and identify its predictors in Sub-Saharan Africa.

## Methods

### Protocol registration and review report

The protocol has been registered with PROSPERO, an international prospective register of systematic reviews (https://www.crd.york.ac.uk/PROSPERO), under registration number CRD42017079375. This meta-analytic review is reported in compliance with the recommendation of Preferred Reporting Items for Systematic Reviews and Meta-Analysis (PRISMA) 2015 statement (21). The PRISMA Explanation and Elaboration document was followed and complemented by A Measurement Tool to Assess Systematic Reviews (AMSTAR-2) tool (22). A PRISMA flow diagram (23) was used to illustrate the article screening and selection process.

### Literature searching

PubMed, EMBASE, CINAHL, Web of Science, African Journals Online and SCOPUS electronic databases were explored to extract all available literatures. Cross-references of included articles and grey literature were also hand searched. In addition, PubMed and SCOPUS cited-by searching of included articles was performed to acquire potentially relevant studies that were potentially missed during database, cross-reference, and grey literature searching. The search strategy (Supplemental Table 1) has been developed in consultation with medical information specialist and Peer Review of Electronic Search Strategies (PRESS) 2015 guideline statement (24).

### Eligibility criteria

The studies were included if they meet the following inclusion criteria: (1) observational (i.e., cross-sectional, case-control, cohort) and (quasi) randomized controlled trial studies; (2) studies conducted in Sub-Saharan Africa between January 2000 to August 2019; and (3) studies published in English. Case reports, case series, expert opinions, qualitative studies, duplicated articles, and studies with substantial incomplete data were excluded.

### Literature screening and selection

Initially, all identified articles were imported into Covidence (25). After duplicate studies were excluded, a pair of reviewers (MA and NB) identified articles by analyzing the abstract and title for relevance to the proposed review topic. Agreement between the reviewers was made by consensus. Then, full-texts were systematically reviewed for further eligibility. Finally, two reviewers (MA and NB) extracted all relevant information, including first author, publication year, country, sample size, study design, prevalence, least adjusted significant predictors, and source of funding using Excel spreadsheet. Disagreement between reviewers was solved through consensus.

### Quality assessment

Two reviewers assessed the quality of selected articles using Newcastle-Ottawa Scale (NOS) for cross-sectional studies (26). The tool has three sections: selection (maximum of 5 stars), comparability (maximum of 5 stars) and outcome (maximum of 5 stars). In this review, studies were ranked as very good if they scored 5 or more stars, good for 4 stars, satisfactory for 3 stars and unsatisfactory for 0-2 stars. Quality assessment and funding sources of the studies are available as a supplementary file (Supplemental Table 2).

### Data analysis

Inverse variance-weighted random-effects model meta-analysis was done to estimate the pooled uptake and odds ratio of predictors with 95% confidence interval. To maintain adequate power, meta-analysis was done if at least five studies were available on a particular outcome of interest. Jackknife sensitivity analysis using the leave-one-out method was used to assess the effect of individual studies on the pooled odds ratio estimate, significance level of estimate and between-study heterogeneity. The study was excluded when the pooled OR estimate increased or decreased by one and changes the significance level after lifting out that particular study from the meta-analysis. Due to small number of studies available for some variables, the change in heterogeneity threshold was not considered as a primary criterion to detect and exclude the outlier study. Narrative synthesis was employed to summarize evidence on predictors. Heterogeneity between studies was tested using Cochran’s Q test and Higgins’s I^2^ test statistic. The risk of publication bias was checked by visualizing funnel plots and Egger’s regression statistical tests. STATA version 11 was used for statistical analysis. To examine the source of heterogeneity, subgroup analysis based on sample size, geographic distribution of the studies and year of publication was carried out.

## Results

### Characteristics of the studies

A total of 3,537 studies were retrieved through database and manual searching. After removing of duplicates (1,577), 93 full-text articles were assessed for further eligibility. Finally, 29 studies with 36,374 women were included in the meta-analysis and qualitative. Only seven studies were included in the meta-analysis for knowledge and cervical cancer screening (figure 1).

**Figure 1.**
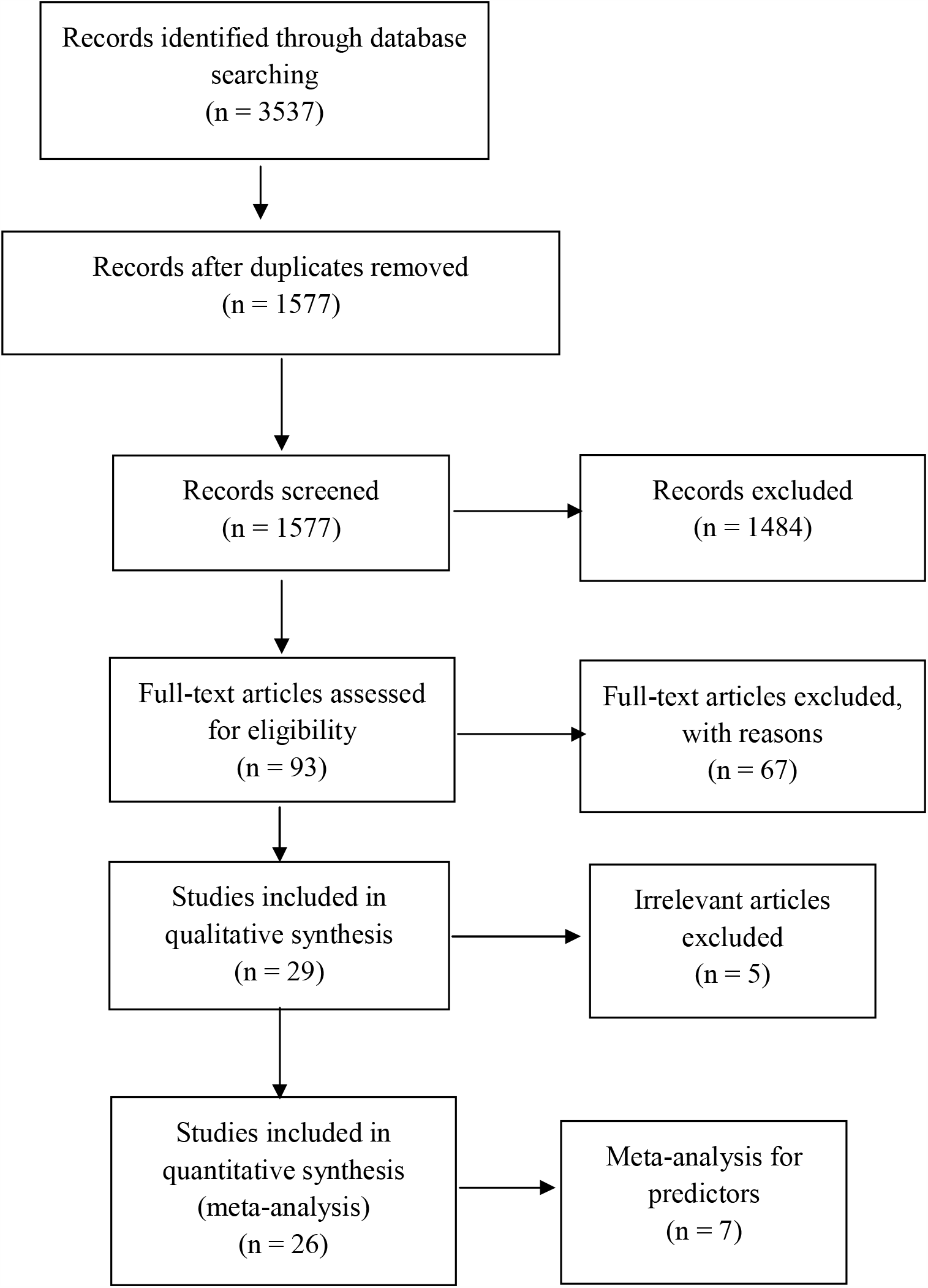
PRISMA flow diagram for predictors of cervical cancer screening, January 2000 to January 2019.

In this review, were included from studies conducted in Sub-Saharan African countries (1 in Ghana, 1 in Burkina Faso, 1 in Botswana, 6 in Nigeria, 7 in Ethiopia, 4 in Kenya, 2 in Uganda, 2 in Tanzania, 2 in Zimbabwe, 1 in Mozambique, 1 in Cameroon and 1 in South Africa). Twenty-eight studies had good quality and one study had good quality score (Table 1).

**Table 1.** Characteristics of the studies in Sub-Saharan Africa, January 2000 to January 2019.

| Study ID | Publication year | Country | Sample size | Screened women | Predictors | Quality score (stars) |
| --- | --- | --- | --- | --- | --- | --- |
| Adanu et.al. (27) | 2010 | Ghana | 3183 | 25 | Lack of formal education<br>Abnormal vaginal bleeding | 7 |
| Sawadego et.al. (28) | 2014 | Burkina Faso | 840 | 93 | Heard about cervical cancer<br>Knowledge about transmission mode<br>Heard about human papillomavirus<br>Oral contraceptive use | 8 |
| Mingo et.al. (29) | 2012 | Botswana | 376 | 271 | Age 31-84<br>Being HIV positive<br>Heard about cervical cancer | 7 |
| CC. Dim et.al. (30) | 2009 | Nigeria | 912 | 82 | Not reported | 5 |
| Chigbu et.al. (31) | 2011 | Nigeria | 3712 | 389 | Not reported | 6 |
| Cunningham et.al. (32) | 2015 | Tanzania | 575 | 35 | Condom use<br>Age 40-49, age >50<br>Health insurance<br>Knowledge about cervical cancer | 7 |
| Tefera et.al. (33) | 2016 | Ethiopia | 634 | 68 | Age 25-35, age 35-49<br>Knowledge about cervical cancer | 8 |
| Aweke et.al. (34) | 2017 | Ethiopia | 595 | 58 | Lack of formal education<br>Primary education<br>Secondary education | 8 |
| Morema et.al. (35) | 2014 | Kenya | 424 | 74 | Lack of awareness about seriousness of disease | 8 |
| Orango'o et.al. (36) | 2016 | Kenya | 2505 | 273 | Being HIV positive<br>Fear of bad result<br>Know place of screening | 8 |
| Tiruneh et.al. (37) | 2017 | Kenya | 9016 | 1750 | Not reported | 8 |
| Lyimo et.al. (38) | 2012 | Kenya | 354 | 80 | Knowledge about cervical cancer | 8 |
| Twinom et.al. (39) | 2015 | Tanzania | 416 | 29 | Not reported | 8 |
| Bayu et.al. (40) | 2016 | Ethiopia | 1286 | 235 | Age 30-39<br>Multiple sexual partners<br>Sexually transmitted diseases<br>Being HIV positive<br>Knowledge about cervical cancer<br>Perceived susceptibility<br>& barriers | 8 |
| Ajibola et.al. (41) | 2016 | Uganda | 338 | 27 | Negative attitude | 8 |
| Olusola et.al. (42) | 2015 | Nigeria | 737 | 110 | Not reported | 5 |
| Akinyemiju et.al. (43) | 2015 | Nigeria | 1236 | 274 | Female provider | 8 |
| Ahmed et.al. | 2016 | South Africa | 500 | 79 | Not reported | 6 |
| Ndejjo et.al. (44) | 2016 | Uganda | 845 | 43 | Getting reproductive care<br>at government facility<br>Know place of screening<br>Ease of getting<br>reproductive service | 8 |
| Sylvia et.al. (45) | 2011 | Zimbabwe | 700 | 63 | Knowledge of screening | 8 |
| Nwankwo et.al. (46) | 2011 | Nigeria | 845 | 36 | Not reported | 7 |
| Bante et.al (47) | 2019 | Ethiopia | 517 | 108 | Age<br>Counseling<br>Positive attitude<br>Visited health facility<br>STIs | 8 |
| Brandao et.al (48) | 2018 | Mozambique | 3177 | 96 | Not reported | 9 |
| Donatus et.al (49) | 2019 | Cameroon | 253 | 110 | Not reported | 4 |
| Gebregzibher et .al (50) | 2019 | Ethiopia | 344 | 59 | Sexual experience<br>Marital status<br>Place of birth<br>Year of study | 7 |
| Getachew et.al (51) | 2019 | Ethiopia | 520 | 130 | Not reported | 8 |
| Ifemelumma et. al (52) | 2019 | Nigeria | 388 | 80 | Not reported | 6 |
| Makuriofa et.al (53) | 2019 | Zimbabwe | 409 | 15 | Not reported | 7 |
| Nigussie et.al (54) | 2019 | Ethiopia | 737 | 114 | Government employee<br>Know someone screened<br>History of gynecologic<br>exam<br>Gender of physician<br>Counseling<br>Knowledge<br>Perceived susceptibility | 8 |

### Uptake of cervical cancer screening

The pooled uptake of cervical cancer screening in Sub-Saharan Africa was 12.87% (95% CI: 10.20, 15.54). There was considerable heterogeneity (I^2^=98.5%), a random effects model was employed (Figure 2), and subgroup analysis was conducted by region, sample size and year of publication. Based on the subgroup analysis, screening uptake ranged from 7.65% in the southern Sub-Saharan African countries to 14.13% in the eastern countries (Figure 3). By sample size, 13.83% of women had screening in a sample size group of less than 800, while 11.34% had screening in studies with sample sizes greater than 800 (Figure 4). Additionally, 13.5% of women were screened among studies published after 2015 (Figure 5). Sensitivity analysis was done; no significant change was noted in the overall odds ratio. There was publication bias, as evidenced by Egger’s test (0.048).

**Figure 2.**
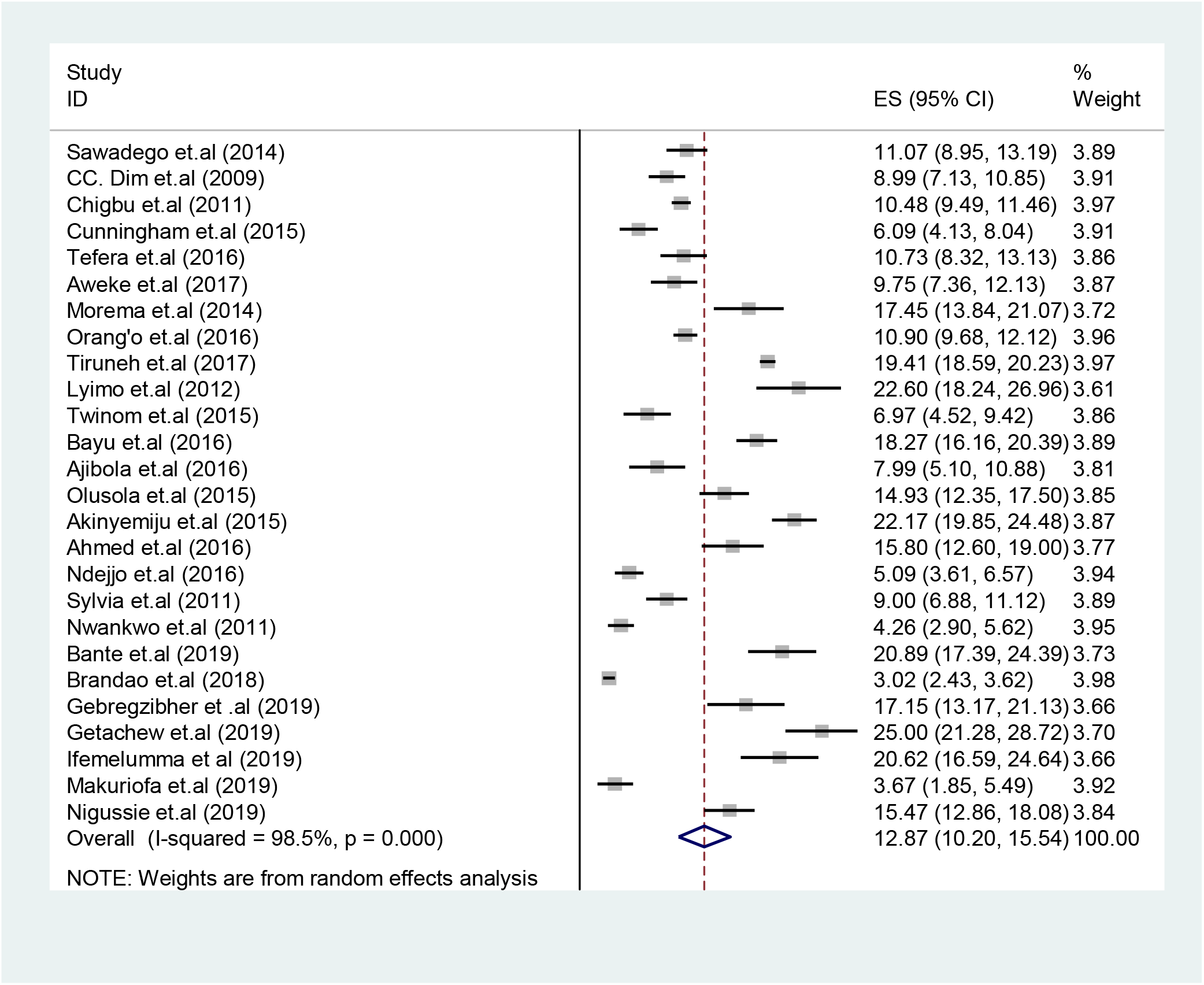
Forest plot of pooled prevalence of cervical cancer screening in Sub-Saharan Africa, January 2000 to January 2019.

**Figure 3.**
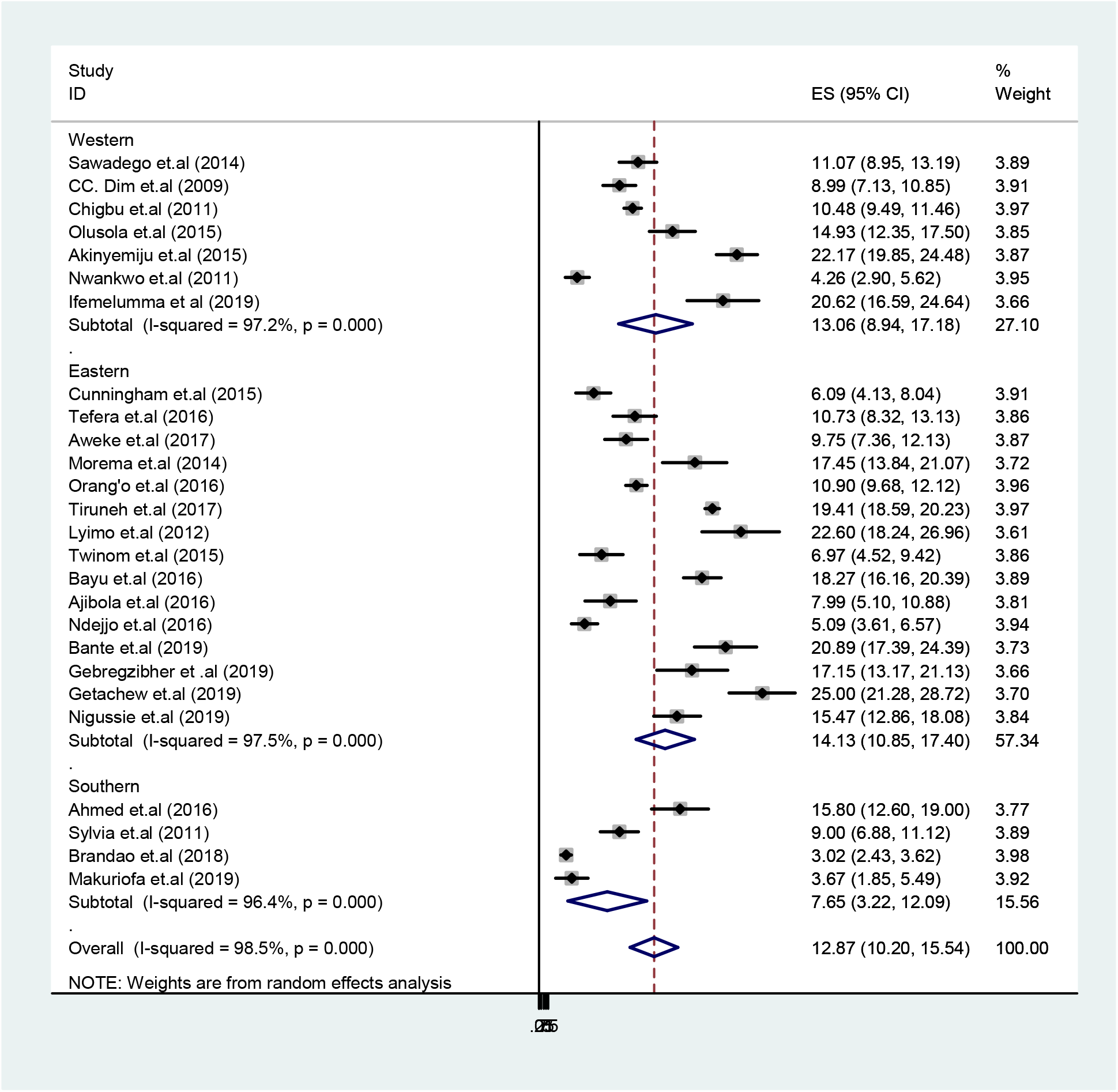
Subgroup analysis by region for uptake of cervical cancer screening in Sub-Saharan Africa from January 2000 to January 2019.

**Figure 4.**
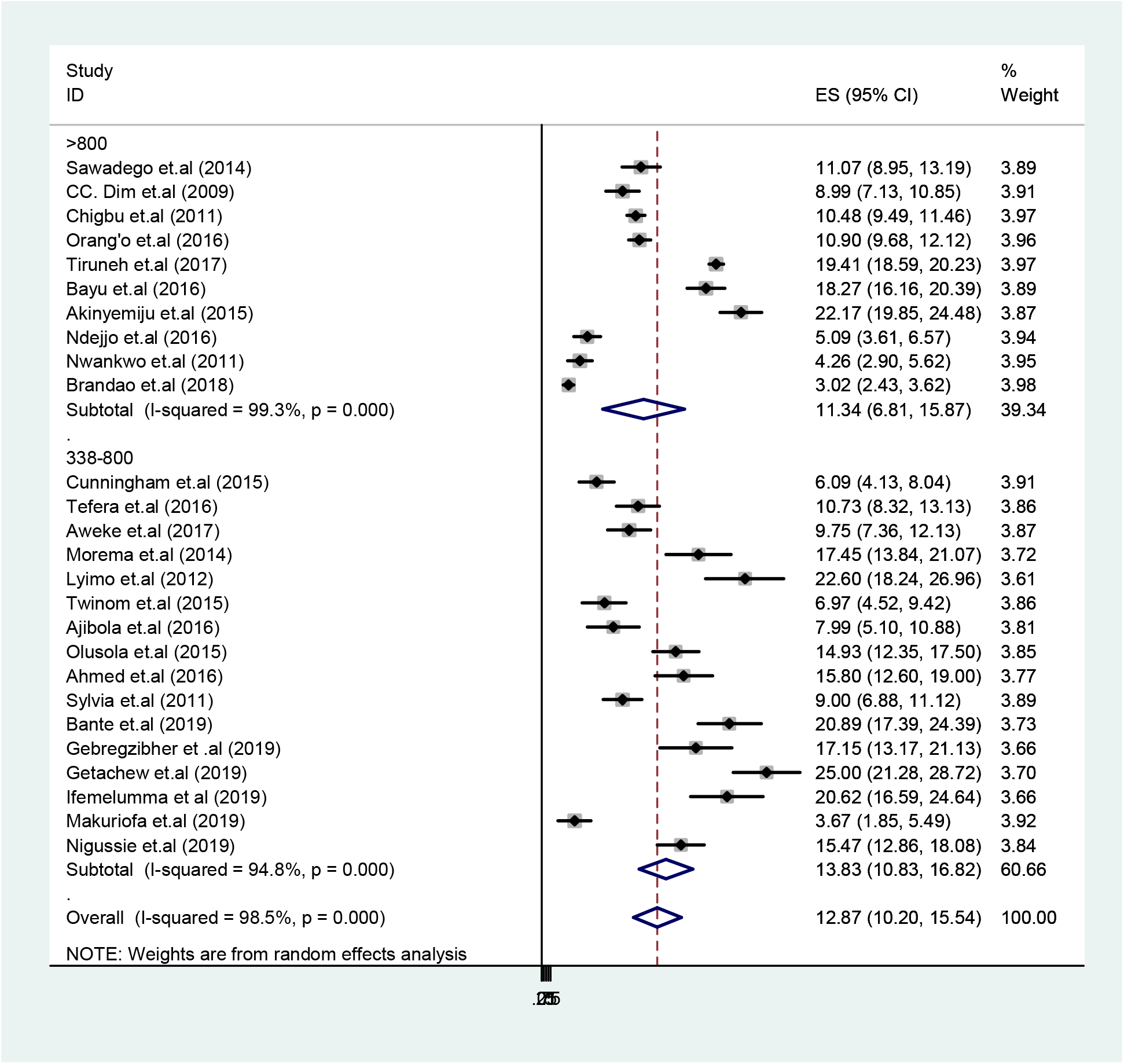
Subgroup analysis by sample size for uptake of cervical cancer screening in Sub-Saharan Africa from January 2000 to January 2019.

**Figure 5.**
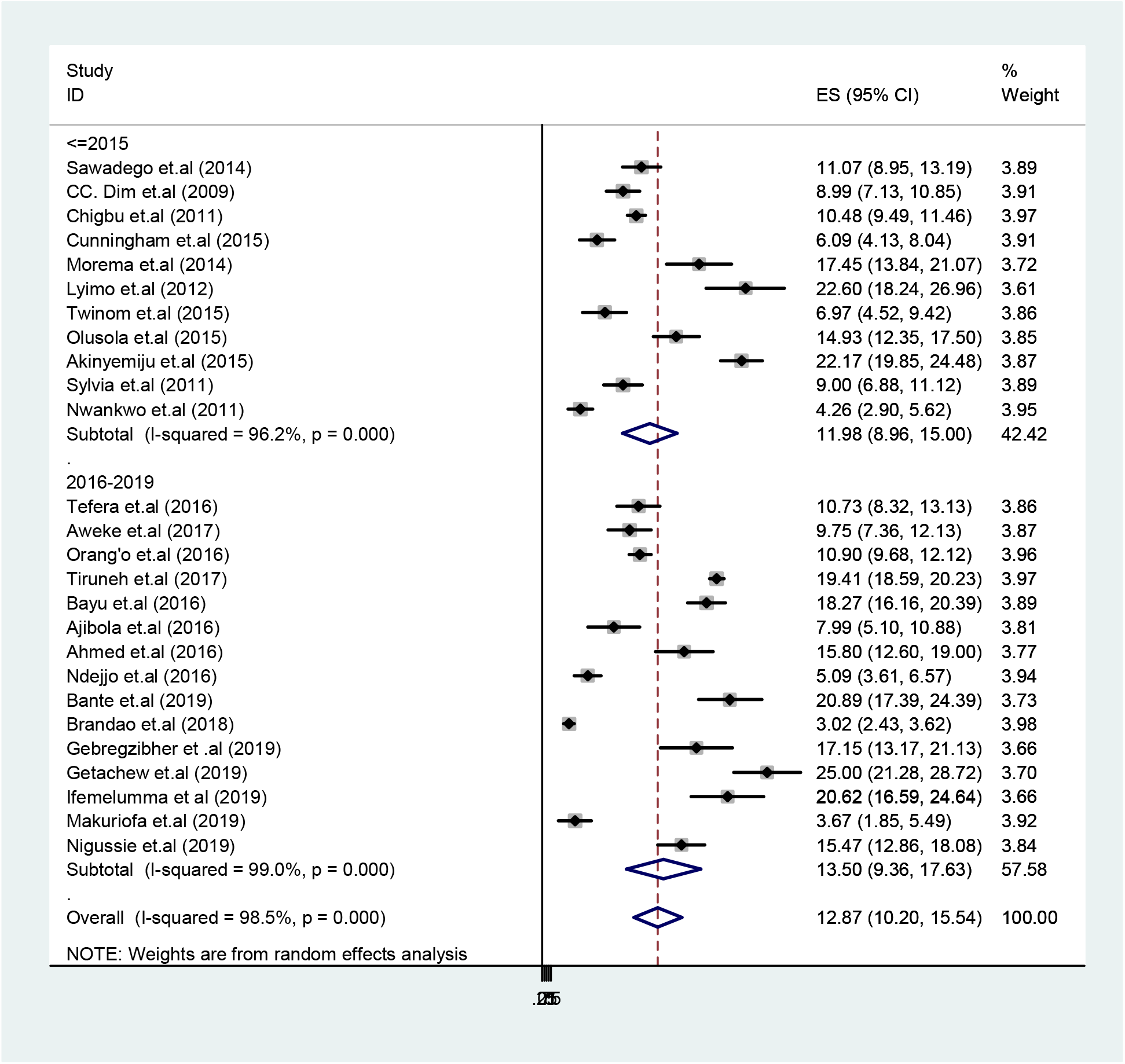
Subgroup analysis by year of publication for uptake of cervical cancer screening in Sub-Saharan Africa from January 2000 to January 2019.

### Predictors of cervical cancer screening

Two studies in Ghana and Ethiopia showed that lack of formal education was significantly associated with low utilization of cervical cancer screening service (27, 34). On the other hand, three studies (29, 36, 40) in the region revealed being HIV positive as a significant predictor for utilization of the screening service. Awareness of place of screening also increased screening uptake in Kenya and Sudan (36, 44). An increase in cervical cancer screening was noted as age increases (47). Tefera and associates (33) reported higher proportion of screened mothers at the age of 25 through 49. Similarly, Bayu and colleagues (29, 32, 40) reported higher utilization of the service with advancement of age (Table 1).

Moreover, negative attitude and perceived susceptibility & barriers lowers the odds of cervical cancer screening uptake (40, 41, 54). Indeed, positive attitude increased service utilization in Ethiopia (47). Akinyemiju and colleagues in Nigeria reported that women tend to be screened when the provider’s gender is female (43). On the contrary, not preferring gender of physician increased screening among Ethiopian Women (54). Two studies (47, 54) in Ethiopia reported counseling about screening were associated with uptake of the service. Abnormal vaginal bleeding (27), heard about HPV & oral contraceptive use (28), health insurance & condom use (32), lack of awareness about seriousness of cervical cancer (35), fear of bad result after screening (36), multiple sexual partners & sexually transmitted diseases (40, 47) and service at government health institutions (44) were also significantly associated with cervical cancer screening uptake (Table 1).

A meta-analysis of seven studies (28, 29, 32, 33, 38, 40, 54) revealed knowledge about cervical cancer screening was significantly associated with cervical cancer screening (OR: 4.81; 95% CI: 3.07, 7.51). There was moderate heterogeneity (I^2^=47.8%), hence random effect model was employed (Figure 6). The Egger’s test (p=0.44) showed no publication bias existed.

**Figure 6.**
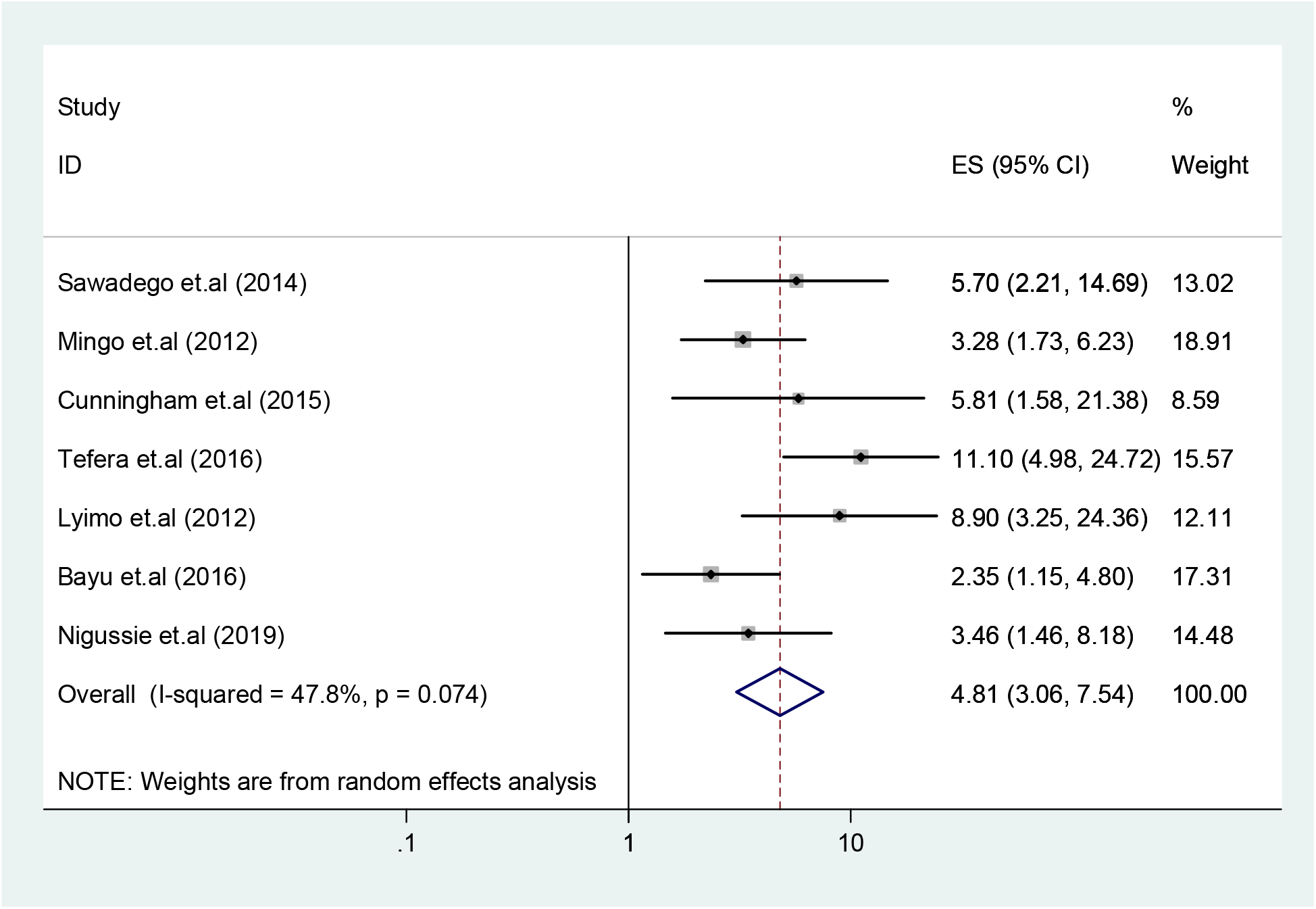
Forest plot for knowledge about cervical cancer screening and uptake of service in Sub Saharan Africa from January 2000 to January 2019.

## Discussion

In this systematic review and meta-analysis, overall uptake of cervical cancer screening was pooled from 26 studies in Sub-Saharan Africa. In addition, significant predictors of cervical cancer screening were also identified. The findings of this review revealed valuable evidence to improve policies and practices aimed at addressing utilization of cervical cancer screening service across the region.

The pooled prevalence of cervical cancer screening in Sub-Saharan Africa was 12.12% (95% confidence interval: 9.48, 14.76) in the present review. This rate is lower than those reported in studies of Chinese-Canadian and Malaysian women, which were 57% (55) and 48.9% (56) respectively. Similarly, this rate is lower than those found in a sample of women with limited primary education in Indonesia (33%-60%), in Malaysia (23%), and in Thailand (67.6), but higher than the Philippines (7.7%) and Vietnam (4.9%) (57). However, these figures should be interpreted cautiously, as they are based on the 2000-2001 WHO estimates and may be dated. Previous literature suggests that the lower uptake of screening in SSA may be due to overcrowding and overburden of health care providers at tertiary facilities (58).

A root cause analysis in low-income countries reported that competing incentives among groups with shared interests in the service, suboptimal working conditions, and lack of cervical cancer prevention support in the political structures of the countries were identified as obstacles for successful cervical screening (59). Another study, a Cochrane review of randomized trials, confirmed that invitations to women due for screening (appointments, letters, phone calls, verbal recommendations, prompts and follow-up letters) increased uptake of screening (60). A systematic review in Low and Middle-income Countries (LMIC) revealed telephone reminders or messages led to increase Pap test uptake (61). Scaling-up of screening services to all primary and secondary health facilities and use of trained paramedical staff may be important to increase uptake. Lower utilization of screening services in Sub-Saharan Africa may also signal that political commitment is needed to improve cervical cancer prevention efforts.

The present systematic review revealed that lack of formal education and inadequate awareness about the seriousness of cervical cancer were associated with low utilization of cervical cancer screening. This finding is consistent with a study in India that reported higher incidence of cervical lesions among illiterate women due to their late presentation to health facilities (58). Community mobilization, including use of village health promotors may be important to increase uptake of screening services. In India, rural cancer registries and campaign approach were found to be useful in detect in cervical cancer at the village level (62). Moreover, the current review noted higher utilization of screening among older women, which is consistent with a study conducted in Malaysia (56). This might be due to the fact that older women tend to seek treatment for their age- or hormone-related complaints. In the Netherlands, women aged 40 to 50 years who felt high personal moral obligation had the highest likelihood of screening uptake (63).

Women in the current review tended to have cervical cancer screening when the provider is female. Similarly, a study in Canada revealed that cervical cancer screening was associated with culturally sensitive health care services (55). Together, these findings may imply the need for culturally appropriate care and outreach. Moreover, the current review showed that women tend to underutilized the screening service due to fear of bad results. Evidence shows potential harms of screening, including anxiety related to positive results (64). The present review also identified negative attitude, perceived susceptibility, and perceived barriers as significant factors for screening uptake. As women’s beliefs may contribute to lower uptake of screening (63), intervention strategies should focus on beliefs and attitudes about cervical cancer.

In the current review, women who had knowledge about cervical cancer are nearly five times more likely to utilize cervical cancer screening than those who did not. Studies have shown that awareness about cervical cancer screening is the priority need in resource-limited countries (58). Similarly, general knowledge about Pap tests was associated with cervical cancer screening among Chinese-Canadian women (55). Additionally, the current finding is in line with a study conducted in Malaysia (56) and systematic reviews in LMIC (65, 66). Awareness about screening services might change the attitude of women to utilize the service. The role of community health workers on educating the community and raising awareness (67) needed to be underscored.

As a limitation, this finding might be prone to risk of bias due to substantial heterogeneity of studies included from different locations. Additionally, differences in cervical screening modalities across the included studies might affect the results of this review.

## Conclusion

Cervical cancer screening uptake is low in Sub-Saharan Africa. Knowledge about cervical cancer was significantly associated with screening. Additionally, education, age, awareness about screening location, HIV-sero status, attitude, provider gender, having heard about HPV, oral contraceptive use, health insurance, condom use, fear of bad result, lack of awareness about seriousness of the disease, multiple sexual partners, sexually transmitted diseases, counseling and receiving service at public institutions were important predictors of cervical cancer screening uptake in the region. Community-based education tailored to culture, literacy level, and pervasive attitudes is recommended to improve uptake of screening.

## Data Availability

The analysis is based on previously published articles.

## Acknowledgments

We are grateful for all authors of the original articles.

## Competing interest

The authors declare no conflict of interest exists.

## Notes

### Competing Interest Statement

The authors have declared no competing interest.

### Clinical Trial

It is a meta-analysis and the protocol has been registered at PROSPERO; CRD42017079375

### Funding Statement

Not applicable.

