## Supplemental Table 1 for "Cervical cancer screening uptake in Sub-Saharan Africa: a systematic review and meta-analysis"

**Table 1**. Search string for “Cervical cancer screening uptake in Sub-Saharan Africa: a systematic review and meta-analysis”

| Component |  | n hits |
| --- | --- | --- |
| 1. | (Cervical cancer[MeSH] OR Cervical Neoplasm*[tiab] OR Cervix Neoplasm*[tiab] OR Cervix Ca*[tiab] OR Cervical Ca* [tiab] OR "Screening uptake"[tiab] OR "Screening utilization"[tiab]) |  |
| 2. | (Screening[MeSH] OR Early Detection[tiab] OR Cancer Screening[tiab] OR Cancer Test*[tiab] OR Early Diagnosis[tiab] OR Cancer Diagnosis[tiab] OR Screenings[tiab]) |  |
| 3. | (“Subsaharan Africa”[tiab] OR Sub-Saharan Africa[tiab] OR Angola[tiab] OR Benin[tiab] OR Botswana[tiab] OR Burkina Faso[tiab] OR Burundi[tiab] OR Cameroon[tiab] OR Cape Verde[tiab] OR Central African Republic[tiab] OR Chad[tiab] OR Comoros[tiab] OR Congo[tiab] OR Côte d'Ivoire[tiab] OR Djibouti[tiab] OR Equatorial Guinea[tiab] OR Eritrea[tiab] OR Ethiopia[tiab] OR Gabon[tiab] OR Gambia[tiab] OR Ghana[tiab] OR Guinea[tiab] OR Guinea-Bissau[tiab] OR Kenya[tiab] OR Lesotho[tiab] OR Liberia[tiab] OR Madagascar[tiab] OR Malawi[tiab] OR Mali[tiab] OR Mauritania[tiab] OR Mauritius[tiab] OR Mozambique[tiab] OR Namibia[tiab] OR Niger[tiab] OR Nigeria[tiab] OR Réunion[tiab] OR Rwanda[tiab] OR Sao Tome[tiab] OR Principe[tiab] OR Senegal[tiab] OR Seychelles[tiab] OR Sierra Leone[tiab] OR Somalia[tiab] OR South Africa[tiab] OR Sudan[tiab] OR Swaziland[tiab] OR Tanzania[tiab] OR Togo[tiab] OR Uganda[tiab] OR Western Sahara[tiab] OR Zambia[tiab] OR Zimbabwe[tiab]) |  |
| 4. | # 1 AND # 2 AND #3 |  |
