## Supplemental Table 2 for "Cervical cancer screening uptake in Sub-Saharan Africa: a systematic review and meta-analysis"

| **Study** | **Source of funding** | **Quality** | | | |
| --- | --- | --- | --- | --- | --- |
|  |  | Selection | Comparability | Outcome | Total score |
| Adanu et.al. 2010 | Not reported | *** | ** | ** | 7 |
| Sawadego et.al. 2014 | Non-governmental | **** | ** | ** | 8 |
| Mingo et.al. 2012 | Non-governmental | *** | ** | ** | 7 |
| Dim CC et.al. 2009 | Not reported | **** |  | * | 5 |
| Chigbu et.al. 2011 | Not reported | **** | * | * | 6 |
| Cunningham et.al. 2015 | Non-governmental | *** | ** | ** | 7 |
| Tefera et.al. 2016 | Not reported | **** | ** | ** | 8 |
| Aweke et.al. 2017 | University/college | **** | ** | ** | 8 |
| Morema et.al. 2014 | Not reported | **** | ** | ** | 8 |
| Orango’o et.al. 2016 | Non-governmental | **** | ** | ** | 8 |
| Tiruneh et.al. 2017 | None | **** | ** | ** | 8 |
| Lyimo et.al. 2012 | Not reported | **** | ** | ** | 8 |
| Twinom et.al. 2015 | Not reported | **** | ** | ** | 8 |
| Bayu et.al. 2016 | University/college | **** | ** | ** | 8 |
| Ajibola et.al. 2016 | Not reported | *** | ** | ** | 8 |
| Olusola et.al. 2015 | Not reported | *** | * | * | 5 |
| Akinyemiju et.al. 2015 | Governmental | **** | ** | ** | 8 |
| Ahmed et.al. 2016 | Not reported | **** | * | * | 6 |
| Ndejjo et.al. 2016 | Non-governmental | **** | ** | ** | 8 |
| Sylvia et.al. 2011 | University/college | **** | ** | ** | 8 |
| Nwankwo et.al. 2011 | Not reported | ***** | * | * | 7 |
| Bante et.al | University/college | **** | ** | ** | 8 |
| Brandao et.al | Governmental | ***** | ** | ** | 9 |
| Donates et.al | Not reported | *** | - | * | 4 |
| Gebregziabher et.al | None | *** | ** | ** | 7 |
| Getachew et.al | University/college | **** | ** | ** | 8 |
| Ifemelumma et.al | None | *** | * | ** | 6 |
| Macurirofa et.al | Non-governmental | **** | * | ** | 7 |
| Nigussie et.al | University/college | **** | ** | ** | 8 |
